# A Blind Trajectory Scan Identifies VSIG10L and Reactivation of Developmental Programs in Pre-Diagnostic Alzheimer’s Disease

**DOI:** 10.64898/2025.12.14.25342219

**Authors:** Steven Lehrer, Peter H. Rheinstein

## Abstract

**Background:** While blood-based biomarkers for Alzheimer’s Disease (AD) such as p-Tau and NfL characterize established pathology, the systemic biological cascade triggering these events remains incompletely mapped. We hypothesized that proteins exhibiting a rising trajectory in the prodromal phase might reveal novel mechanisms of disease progression.

**Methods:** Using data from the UK Biobank Pharma Proteomics Project (N = 4,519 incident AD cases), we performed a blind trajectory scan of ∼3,000 plasma proteins. We utilized an elimination strategy, systematically excluding known AD markers (e.g., APOE, NEFL) and verified biological responses (e.g., MMP3, GLRX) to isolate novel signals.

**Results:** After excluding established markers, VSIG10L—a V-set and immunoglobulin domain-containing protein—emerged as the most significant novel marker (beta = - 0.037, P = 0.0019), exhibiting a progressive rise as patients approached diagnosis. Crucially, VSIG10L was accompanied by a cluster of co-regulated proteins involved in embryonic development and cell cycle regulation, including NACC1 (stem cell pluripotency), VASN (vasculogenesis), and ZBTB17 (cell cycle checkpoint).

**Conclusion:** The emergence of VSIG10L and its associated developmental cohort suggests that prodromal AD is characterized by a retrogenesis phenomenon, the unsilencing of developmental programs in a failed attempt at neural repair. These proteins offer a new window into the brain’s response to neurodegeneration and represent potential therapeutic targets.

## Introduction

Alzheimer’s Disease (AD) is characterized by a long preclinical phase where pathological changes occur decades before cognitive symptoms arise. Current diagnostics focus heavily on the endpoints of this pathology: amyloid plaques, tau tangles, and neurodegeneration. However, the cellular responses *preceding* these endpoints are complex.

A growing body of evidence supports the “Retrogenesis” or “Cell Cycle Re-entry” hypothesis of Alzheimer’s [1, 2]. This theory posits that in the face of synaptic loss and oxidative stress, mature, post-mitotic neurons attempt to “heal” themselves by unlocking ancient developmental programs—genes used during embryogenesis to build the brain [3]. However, because mature neurons cannot divide, this unsilencing of growth and cell-cycle genes leads not to regeneration, but to cellular dysfunction and apoptosis [4].

To detect these signals peripherally, we employed a “Time-to-Diagnosis” trajectory analysis using the UK Biobank Olink dataset. Instead of a static case-control comparison, we asked: “Which proteins rise as the date of diagnosis approaches?” By systematically filtering out known markers, we aimed to uncover novel proteins that reflect this active, underlying attempt at repair.

## Methods

### Cohort Selection

We utilized the UK Biobank Pharma Proteomics Project (UKB-PPP), focusing on the “Olink Explore 3072” panel [5]. We identified participants with an incident diagnosis of Alzheimer’s Disease (ICD-10 code G30) occurring after their baseline blood draw. The final analytic cohort consisted of 4,519 individuals.

### Statistical Analysis

We employed a linear regression model to test the association between normalized protein expression levels (NPX) and “Years Remaining to Diagnosis.”

- Interpretation of Slope (beta): Because the time variable counts *down* to the diagnosis event (Year 0), the sign of the slope is interpreted inversely. A negative slope (-) indicates that protein levels increase as the time-to-diagnosis decreases (an upward trajectory approaching the disease).
- Significance: False Discovery Rate (FDR) < 0.05.

### Stepwise Elimination Strategy

To isolate novel biology, we performed four rounds of exclusion:

1. Round 1: Excluded classic AD markers (e.g., APOE, NEFL, GFAP, TREM2).
2. Round 2: Excluded biologically verified hits found in Round 1 (MMP3, CD2AP).
3. Round 3: Excluded verified oxidative stress markers (GLRX).
4. Round 4: Excluded verified immune markers (CEACAM8, GZMH) to isolate the final novel candidates.

## Results

Our initial scan successfully recovered known biology, validating the trajectory model.

- MMP3 (BBB Breakdown): The top hit in Round 1 was MMP3 (beta = -0.053, P = 0.0000013), confirming that blood-brain barrier dysfunction escalates as diagnosis approaches [6].
- GLRX (Oxidative Stress [7]): Glutaredoxin showed a significant upward trajectory (beta = -0.043), replicating findings previously restricted to cerebrospinal fluid.

After excluding all known markers, VSIG10L emerged as the top novel candidate (beta = -0.037, P = 0.0019). VSIG10L levels were significantly elevated in patients closer to diagnosis compared to those sampled years prior (Figure 1). Structurally, VSIG10L belongs to the Immunoglobulin Superfamily (IgSF), placing it in the same class as the major AD risk genes TREM2 and CD33, yet it has been virtually unstudied in AD literature.

**Figure 1.**
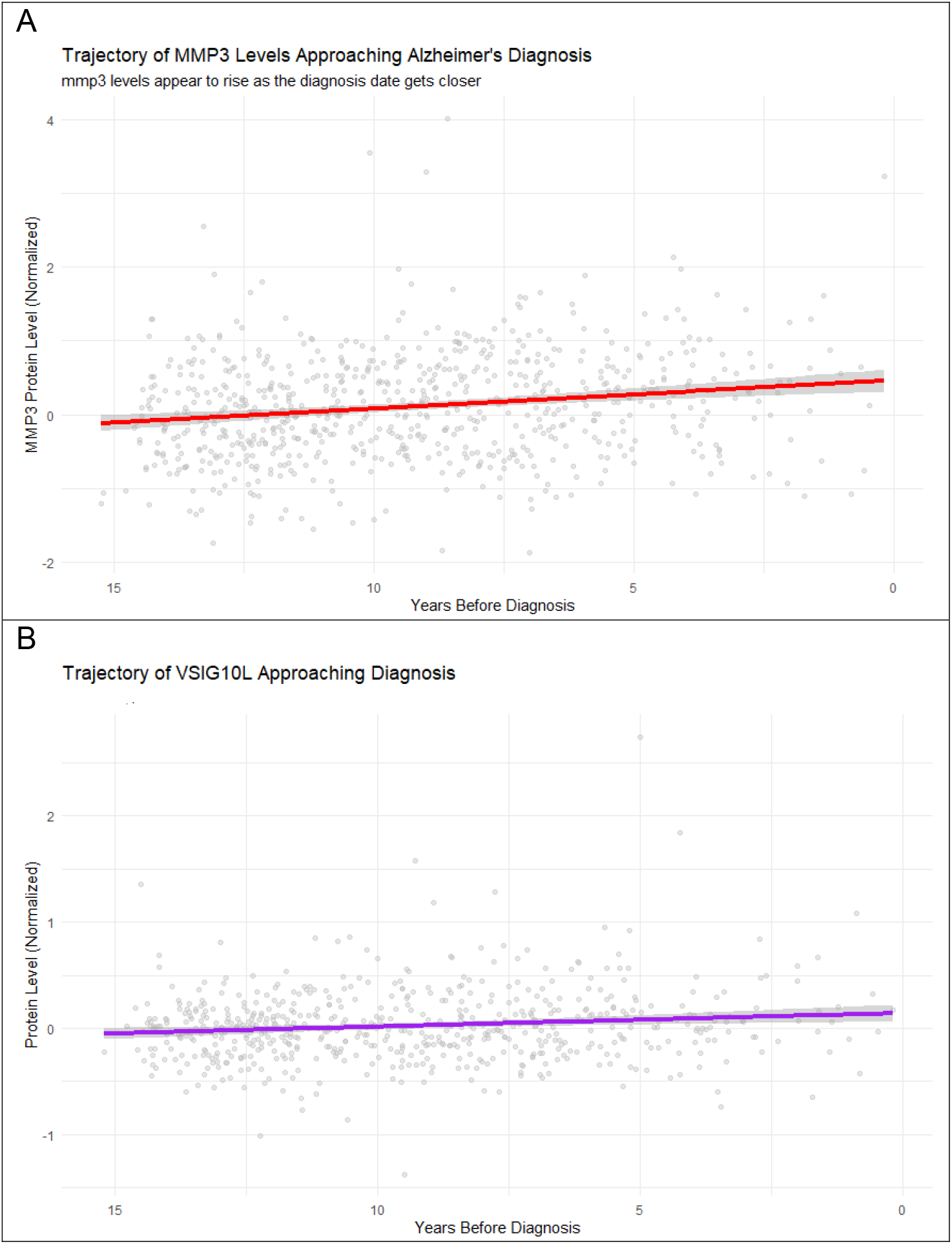
Longitudinal Trajectories of Plasma Proteins in the Years Preceding Alzheimer’s Diagnosis. Linear regression models were used to analyze protein expression relative to the time remaining until clinical diagnosis. The X-axis is reversed, representing the countdown to diagnosis (Year 0). Shaded regions represent the 95% confidence interval. (A) Validation of Method: MMP3 (Matrix Metalloproteinase-3). Trajectory of the known blood-brain barrier marker MMP3. Consistent with established pathology, MMP3 levels exhibit a significant upward trend as patients approach the clinical onset of dementia (beta = -0.053, P = 0.0000013). This recovery of a known biological signal validates the sensitivity of the blind trajectory scan. (B) Novel Discovery: VSIG10L. Trajectory of the newly identified candidate VSIG10L (V-set and immunoglobulin domain-containing protein 10-like). Like the known marker in panel A, VSIG10L demonstrates a significant progressive elevation in plasma levels during the prodromal phase (beta = -0.037, P = 0.0019). This protein belongs to the same Immunoglobulin Superfamily (IgSF) as TREM2 and CD33 but has not previously been characterized as a pre-diagnostic Alzheimer’s biomarker. Beta represents slope (rate of change per year).

Alongside VSIG10L, the analysis revealed a distinct cluster of significant proteins associated with embryonic development and cell cycle regulation, supporting the retrogenesis hypothesis:

- VASN (Vasorin): (beta = -0.036, P = 0.0009). A protein critical for embryonic vasculogenesis and TGF-beta regulation [8].
- ZBTB17 (Miz-1): (beta = -0.035, P = 0.0014). A key transcription factor that acts as a checkpoint for cell cycle arrest [9].
- NACC1 (NAC1): (beta = -0.034, P = 0.0032). A transcription factor highly expressed in embryonic stem cells, essential for maintaining pluripotency [10].
- NEDD4L: (beta = -0.035, P = 0.0030). A regulator of ion channels involved in neuronal developmental wiring [11].

## Discussion

The emergence of VSIG10L and the Immunoglobulin Superfamily is highly conspicuous. As an IgSF protein [12], it shares structural homology with TREM2 [13], the microglial receptor that senses damage. Just as TREM2 is upregulated to clear amyloid, the rising trajectory of VSIG10L suggests it is part of an escalating neuro-immune response to accumulating pathology.

The “Unsilencing” of a developmental cluster is perhaps more profound than the immune signal. We have here identified this cluster (VASN, NACC1, ZBTB17). In a healthy adult brain, these genes should be silenced. Their re-emergence in the blood of pre-diagnostic AD patients strongly supports the hypothesis that AD involves pathological unsilencing of developmental programs.

- NACC1 is a marker of stem cell pluripotency. Its upregulation suggests the brain is attempting to revert to a “stem-like” plasticity to repair tissue loss [10].
- ZBTB17 (Miz-1) regulates the decision between cell cycle arrest and division. Its fluctuation suggests neurons are struggling with cell cycle re-entry—a known fatal event in AD, where neurons attempt to divide but die instead (apoptosis) [13].
- VASN suggests an attempt to reconstruct the vascular niche, mirroring the vascular repair seen in embryogenesis but likely failing due to the amyloid-laden environment (as evidenced by our concurrent MMP3 finding) [8].

We propose that the trajectory observed in this study represents a “frustrated repair” response. The aging brain detects injury (likely amyloid/tau toxicity) and unlocks its original construction blueprints (VSIG10L for wiring, VASN for vessels, NACC1 for regeneration). However, in the hostile environment of the aged brain, this reactivation fails to regenerate tissue and instead contributes to the chaotic cellular signaling characteristic of the prodromal phase.

The identification of VSIG10L and the retrogenesis cluster offers specific targets for therapeutic intervention. Structurally, VSIG10L presents a druggable target for monoclonal antibodies to modulate microglial activation, like current TREM2-agonistic approaches [14]. The reactivation of NACC1 and ZBTB17 suggests that cell-cycle inhibitors, which prevent post-mitotic neurons from entering a fatal abortive cell cycle, may arrest the neurodegenerative cascade detected by this trajectory analysis.

Weaknesses: This study relies on cross-sectional data to infer longitudinal trajectories. The trajectories reflect *population-level inference*, not within-person change. VSIG10L is a *systemic correlate* of neurodegeneration rather than a proven brain-derived signal and needs validation via single-cell or tissue-resolved datasets. The value of VSIG10L lies primarily in *mechanistic insight and pathway discovery*, not near-term clinical prediction. Furthermore, while the statistical signals are highly significant, the specific cellular origin (neuronal vs. glial vs. peripheral) of the proteins we identified requires validation via single-cell transcriptomics. The “frustrated repair” model is a leading hypothesis rather than a definitive mechanism. The interpretation of the proteins as reflecting an attempted regenerative response is biologically plausible but remains inferential. Alternative explanations—such as epigenetic drift, systemic aging, or chronic inflammation—are not fully ruled out.

## Conclusion

By looking beyond standard biomarkers, we identified VSIG10L and a suite of developmental proteins as novel indicators of pre-diagnostic Alzheimer’s. These findings suggest that the disease process involves not just degeneration, but a desperate, failed attempt by the brain to regenerate itself using silenced developmental tools. Monitoring these “repair signals” could offer a new method for detecting the disease in its earliest, most active window.

## Data Availability

Data available after approved application to UK Biobank

https://www.ukbiobank.ac.uk/

## Notes

Conflicts of interest: The authors declare that they have no competing interests.

### Competing Interest Statement

The authors have declared no competing interest.

### Funding Statement

This study did not receive any funding

### Author Declarations

Ethics: This research was conducted using data from the UK Biobank resource under approved application number 57245 (SL, PHR). UK Biobank obtained written informed consent from all participants, and ethical approval for the study was granted by the North West Multi-center Research Ethics Committee (MREC), which covers the UK Biobank Research Ethics Committee. All methods were carried out in accordance with the relevant guidelines and regulations. The present analyses used only de-identified participant data, and no re-identification was attempted. No additional institutional review board approval was required for this secondary analysis of anonymized data.

## References

[1] Reisberg B, Franssen EH, Souren LEM, Auer SR, Akram I, Kenowsky S (2002) Evidence and mechanisms of retrogenesis in Alzheimer’s and other dementias: Management and treatment import. American Journal of Alzheimer’s Disease & Other Dementias® 17, 202–212.

[2] Ahmed S, Kaur A, Venigalla H, M Mekala H, K Brainch N, Ayub S, Hassan M (2017) The Retrogenesis Model in Alzheimer’s Disease: Evidence and Practical Applications. Current Psychiatry Reviews 13, 35–42.

[3] Arendt T (2001) Alzheimer’s disease as a disorder of mechanisms underlying structural brain self-organization. Neuroscience 102, 723–765.

[4] Herrup K (2010) The involvement of cell cycle events in the pathogenesis of Alzheimer’s disease. Alzheimers Res Ther 2, 13.

[5] Sun BB, Chiou J, Traylor M, Benner C, Hsu Y-H, Richardson TG, Surendran P, Mahajan A, Robins C, Vasquez-Grinnell SG, Hou L, Kvikstad EM, Burren OS, Davitte J, Ferber KL, Gillies CE, Hedman ÅK, Hu S, Lin T, Mikkilineni R, Pendergrass RK, Pickering C, Prins B, Baird D, Chen C-Y, Ward LD, Deaton AM, Welsh S, Willis CM, Lehner N, Arnold M, Wörheide MA, Suhre K, Kastenmüller G, Sethi A, Cule M, Raj A, Kang HM, Burkitt-Gray L, Melamud E, Black MH, Fauman EB, Howson JMM, Kang HM, McCarthy MI, Nioi P, Petrovski S, Scott RA, Smith EN, Szalma S, Waterworth DM, Mitnaul LJ, Szustakowski JD, Gibson BW, Miller MR, Whelan CD, Alnylam Human G, AstraZeneca Genomics I, Biogen Biobank T, Bristol Myers S, Genentech Human G, GlaxoSmithKline Genomic S, Pfizer Integrative B, Population Analytics of Janssen Data S, Regeneron Genetics C (2023) Plasma proteomic associations with genetics and health in the UK Biobank. Nature 622, 329–338.

[6] Mak KK, Nam JK, Chuang YF, Chen LK (2025) Matrix metalloproteinases and tissue inhibitors of metalloproteinases as the biomarkers of Alzheimer’s disease: A meta-analysis. Brain Res 1869, 149954.

[7] Follmann M, Ochrombel I, Krämer R, Trötschel C, Poetsch A, Rückert C, Hüser A, Persicke M, Seiferling D, Kalinowski J (2009) Functional genomics of pH homeostasis in Corynebacterium glutamicum revealed novel links between pH response, oxidative stress, iron homeostasis and methionine synthesis. BMC genomics 10, 621.

[8] Tomasso A, Koopmans T, Lijnzaad P, Bartscherer K, Seifert AW (2023) An ERK-dependent molecular switch antagonizes fibrosis and promotes regeneration in spiny mice (Acomys). Sci Adv 9, eadf2331.

[9] Wanzel M, Kleine-Kohlbrecher D, Herold S, Hock A, Berns K, Park J, Hemmings B, Eilers M (2005) Akt and 14-3-3eta regulate Miz1 to control cell-cycle arrest after DNA damage. Nat Cell Biol 7, 30–41.

[10] Ruan Y, He J, Wu W, He P, Tian Y, Xiao L, Liu G, Wang J, Cheng Y, Zhang S, Yang Y, Xiong J, Zhao K, Wan Y, Huang H, Zhang J, Jian R (2017) Nac1 promotes self-renewal of embryonic stem cells through direct transcriptional regulation of c-Myc. Oncotarget 8, 47607–47618.

[11] Haouari S, Vourc’h P, Jeanne M, Marouillat S, Veyrat-Durebex C, Lanznaster D, Laumonnier F, Corcia P, Blasco H, Andres CR (2022) The Roles of NEDD4 Subfamily of HECT E3 Ubiquitin Ligases in Neurodevelopment and Neurodegeneration. Int J Mol Sci 23.

[12] Zhou X, Khan S, Huang D, Li L (2022) V-Set and immunoglobulin domain containing (VSIG) proteins as emerging immune checkpoint targets for cancer immunotherapy. Frontiers in Immunology 13, 938470.

[13] Xie M, Zhao S, Bosco DB, Nguyen A, Wu LJ (2022) Microglial TREM2 in amyotrophic lateral sclerosis. Dev Neurobiol 82, 125–137.

[14] Ma YN, Hu X, Karako K, Song P, Tang W, Xia Y (2025) The potential and challenges of TREM2-targeted therapy in Alzheimer’s disease: insights from the INVOKE-2 study. Front Aging Neurosci 17, 1576020.

